# COVID-19-related school disruptions and well-being of children and adolescents in Geneva

**DOI:** 10.1101/2022.01.07.21268224

**Authors:** Viviane Richard, Roxane Dumont, Elsa Lorthe, Hélène Baysson, María-Eugenia Zaballa, Rémy P. Barbe, Klara M. Posfay-Barbe, Idris Guessous, Silvia Stringhini

## Abstract

**Background:** Various studies showed the negative impact of COVID-19-related lockdowns and school closures on the well-being of children and adolescents. However, the prevalence and consequences of occasional short-term school disruptions due to COVID-19-related quarantine or isolation remain unknown. This study evaluated their impact on the well-being and stress level of children and adolescents.

**Methods:** In June/July 2021, we conducted a survey selecting a representative sample of children and adolescents of a Swiss canton population. Parents of school-aged children reported information about them missing school because of COVID-19, from August 2020 to June 2021, as well as about their health-related quality of life (HRQoL) measured with the KINDL® scale and their stress level.

**Results:** Among the 538 participants, 216/538 (40.1%) pupils missed school at least once for COVID-19-related causes, with a total of 272 absences. We observed no relationship between the frequency of COVID-19-related absences and the HRQoL or stress level, even when stratifying by the type of absence or socio-demographic factors.

**Discussion:** Overall, these findings are reassuring in that quarantines and related school disruptions, which we know are a common and effective way of controlling SARS-CoV-2 transmission, did not seem to meaningfully impact children and adolescent’s wellbeing and stress. Finding the right balance between SARS-CoV-2 control and young populations’ well-being is challenging, and the current results provide additional information for decision makers.

## Background

Various studies showed the negative impact of COVID-19-related lockdowns and school closures on the well-being of children and adolescents [1–4]. In Switzerland, schools were closed between March and May/June 2020 [5] and remained open thereafter. Since the reopening, pupils were required to isolate if infected by SARS-CoV-2 and to quarantine in case of close contact with an infected person. Over the study period, the local COVID-19 incidence was high and quarantines of entire class were also possible in case of a significant cluster.

The prevalence and consequences of these occasional short-term school disruptions due to COVID-19 remain unknown. This study evaluated their impact on the well-being and stress level of children and adolescents.

## Methods

In June/July 2021, we conducted a survey selecting a representative sample of children and adolescents of a Swiss canton population (Ethics Committee ID: 2020-0088) [6]. Parents of school-aged children reported information about them missing school because of COVID-19, from August 2020 to June 2021. Parents also completed questionnaires about their child(ren)’s sex, school level, health-related quality of life (HRQoL) measured with the KINDL® scale and stress level, as well as household financial situation.

## Results

The sample comprised 538 participants (258 [48.0%] are female) from 4 to 18 years old (Mean: 10.6; SD: 3.6).

From August 2020 to June 2021, 216/538 (40.1%) pupils missed school at least once for COVID-19-related causes: 174/538 (32.3%) missed school once, 37/538 (6.8%) two or three times and 5/538 (0.9%) four or five times, with a total of 272 absences. 179/538 (33.3%) pupils missed school because of isolation or quarantine while the class remained open, 23/538 (4.3%) because the class or institution closed due to COVID-19 and 14/538 (2.6%) for both reasons. Most of the absences 179/272 (65.8%) lasted more than a week.

We observed no relationship between the frequency of COVID-19-related absences and the HRQoL (Table 1) or stress level (Supplementary 1), even when stratifying by the type of absence or socio-demographic factors.

**Table 1.** Health-Related Quality of Life (HRQoL) of children and adolescents according to the number of school disruptions, stratified by the type of absence and socio-demographic characteristics

|  |  | Absence from school |  |  | p-value<sup>b</sup> |
| --- | --- | --- | --- | --- | --- |
|  |  | None | Once | Twice or more |  |
|  | N | Mean (SD)<sup>a</sup> | Mean (SD)<sup>a</sup> | Mean (SD)<sup>a</sup> |  |
| **Total** | 538 | 82.1 (8.4) | 81.1 (9.3) | 82.7 (9.3) | 0.36 |
| **School/class closure** | 538 | 82.0 (8.6) | 79.5 (10.7) | - | 0.19 |
| **Individual absence** | 538 | 81.8 (8.7) | 81.8 (8.7) | 83.0 (10.0) | 0.46 |
| **Sex** |  |  |  |  |  |
| Female | 258 | 81.1 (8.8) | 81.5 (10.4) | 81.6 (12.2) | 0.26 |
| Male | 273 | 83.0 (7.8) | 80.7 (8.3) | 84.0 (6.4) | 0.75 |
| Diverse | 7 | 87.5 (8.3) | 82.5 (7.1) | 79.2 (9.4) | 0.26 |
| **School level** |  |  |  |  |  |
| Primary (4-12 years old) | 347 | 83.9 (7.9) | 82.4 (8.8) | 83.6 (9.4) | 0.26 |
| Lower secondary (12-15 years old) | 121 | 79.0 (8.4) | 79.0 (8.8) | 81.3 (7.7) | 0.75 |
| Upper secondary (15-18 years old) | 64 | 78.4 (7.2) | 79.7 (12.1) | 80.0 (11.5) | 0.26 |
| **Household financial situation<sup>c</sup>** |  |  |  |  |  |
| Very good | 182 | 83.4 (7.5) | 83.9 (7.9) | 81.8 (10.1) | 0.63 |
| Good | 189 | 82.0 (8.3) | 77.5 (8.8) | 83.7 (5.3) | 0.003 |
| Medium to poor | 107 | 79.3 (10.2) | 80.6 (9.7) | 82.6 (12.7) | 0.32 |
| No answer | 55 | 83.5 (6.2) | 84.0 (10.3) | 84.7 (5.5) | 0.81 |

<sup>a</sup> HRQoL reported by parents with the KINDL<sup>®</sup> scale
<sup>b</sup> Kruskal-Wallis test; as most p-values were higher than 0.1, we considered that an adjustment for multiple comparisons was not needed.
<sup>c</sup> Very good: affluent, can save money; Good: earning sufficient to cover expenses and to face minor unexpected expenses; Medium to poor: cannot face unexpected expenses and/or need external financial support

As school- and friends-related well-being could be more specifically affected by school disruptions, sensitivity analyses were performed with the corresponding subscales of the KINDL®; results were similar to those reported for the whole scale (available from the authors).

## Discussion

About 40% of children and adolescents missed school at least once because of COVID-19 over the school year 2020-2021. Findings suggest that their well-being and stress level were not affected by these occasional disruptions, even when stratifying by type of absence and socio-demographic factors. To the best of our knowledge, this study is the first to examine school disruptions due to COVID-19 outside a general lockdown.

The differences in duration and context may explain the discrepancy with findings from lockdowns of early stages of the pandemic [1–4]. In spring 2020, school closures lasted on average 11 weeks in Europe [7] and were characterised by an uncertain and worrying environment. Subsequent COVID-19-related isolations or quarantines implying school disruption were shorter, lasting 7 to 10 days. Moreover, most children were only affected once over the school year. Therefore, short-term school disruptions due to COVID-19 may be assimilated to school absenteeism due to “usual infections”, which is commonly reported to range between 4.5 [8] to 7.6 [9] school days per year.

Additionally, pandemic-related sanitary measures may have decreased the transmission of other common infections, so that overall pupils might have missed the same number of, or even fewer school days in the current pandemic situation than in a normal school year. More studies are needed to better understand how COVID-19 shaped school absenteeism.

The major limitation of this study is the fact that we only examined HRQoL and stress level; we could not measure potential educational losses, nor the impact of online teaching. Furthermore, data is self-reported with a limited sample size.

Overall, these findings are reassuring in that quarantines and related school disruptions, which we know are a common and effective way of controlling SARS-CoV-2 transmission, did not seem to meaningfully impact children and adolescent’s wellbeing and stress. However, school absences should not be trivialized as they are linked to poorer educational achievements [10]. Finding the right balance between SARS-CoV-2 control and young populations’ well-being is challenging, and the current results provide additional information for decision makers.

## Data Availability

All data produced in the present study are available upon reasonable request to the authors

## Acknowledgments

We are grateful to the Specchio-COVID19 group whose members are listed below, as well as to all the participants whose contributions were invaluable to the study.

