## Supplementary 1 for "COVID-19-related school disruptions and well-being of children and adolescents in Geneva"

**Supplementary 1.** Stress experienced by children and adolescents according to the number of school absences, stratified by the type of absence and socio-demographic characteristics

| | | Absence from school | | | |
|---|---|---|---|---|---|
| | | None | Once | Twice or more | |
| | N | Mean (SD)[a] | Mean (SD)[a] | Mean (SD)[a] | p-value[b] |
| **Total** | 538 | 2.1 (1.2) | 2.2 (1.2) | 2.2 (1.2) | 0.31 |
| **School/class closure** | 538 | 2.1 (1.2) | 2.4 (1.3) | - | 0.28 |
| **Individual absence** | 538 | 2.1 (1.2) | 2.2 (1.2) | 2.3 (1.3) | 0.57 |
| **Sex** | | | | | |
| Female | 258 | 2.3 (1.2) | 2.3 (1.3) | 2.2 (1.3) | 0.39 |
| Male | 273 | 1.9 (1.1) | 2.2 (1.1) | 2.2 (1.2) | 0.39 |
| Diverse | 7 | 2.0 (1.0) | 1.5 (0.7) | 2.5 (2.1) | 0.82 |
| **School level** | | | | | |
| Primary (4-12 years old) | 347 | 2.0 (1.2) | 2.1 (1.2) | 2.2 (1.3) | 0.39 |
| Lower secondary (12-15 years old) | 121 | 2.3 (1.2) | 2.5 (1.2) | 2.0 (1.2) | 0.39 |
| Upper secondary (15-18 years old) | 64 | 2.4 (1.2) | 2.4 (1.4) | 2.5 (1.0) | 0.91 |
| **Household financial situation[c]** | | | | | |
| Very good | 182 | 1.9 (1.0) | 2.1 (1.2) | 2.2 (1.3) | 0.33 |
| Good | 189 | 2.1 (1.2) | 2.4 (1.1) | 1.9 (1.0) | 0.23 |
| Medium to poor | 107 | 2.5 (1.5) | 2.5 (1.4) | 2.5 (1.4) | 0.95 |
| No answer | 55 | 2.1 (1.0) | 2.0 (1.2) | 2.3 (1.2) | 0.62 |

[a] Stress level reported by parents answering the question: Currently, how would you assess your child's stress level on a scale from 1 (lowest) to 6 (highest)?

[b] Kruskal-Wallis test; as p-values were higher than 0.1, we considered that an adjustment for multiple comparisons was not needed.

[c] Very good: affluent, can save money; Good: earning sufficient to cover expenses and to face minor unexpected expenses; Medium to poor: cannot face unexpected expenses and/or need external financial support
